# High-dimensional Characterization of Genome–Environment Fitness Landscapes in *Klebsiella pneumoniae*

**DOI:** 10.64898/2026.05.28.26354339

**Authors:** Ge Zhou, Georgia Williams, Mathew T. Millner, Raneem AlHirayban, Wejdan Alosaimi, Omniya Fallatah, Adam J Hart, Mohammed Malaikah, Sara Iftikhar, Huda Ahmad, Mohammad Roghanian, Ville Mustonen, Rfeef AlYami, Manuel Banzhaf, Danesh Moradigaravand

## Abstract

**Background:** Bacterial fitness is shaped by interactions between genome variation and environmental context, yet how these interactions determine its predictability and heritability remains unclear. In the clinically important pathogens of *Klebsiella pneumoniae*, a leading cause of hospital-acquired infections, this question is particularly pressing. Despite extensive genomic characterization, we still lack a systematic understanding of how genome-wide variation translates into fitness across diverse environments in *K. pneumoniae*.

**Methods:** We filled this gap by profiling a systematic collection of 1,462 clinical *K. pneumoniae* isolates across 214 diverse environmental and pharmacological stress conditions using high-throughput chemical genomics. Fitness was quantified from colony growth and integrated with whole-genome sequencing data. Genome-wide association analyses identified genetic determinants of fitness, and machine learning models incorporating genomic features were used to predict fitness.

**Results:** Fitness exhibited a strongly environment-dependent genetic architecture, with modest but significant concordance between genetic background and phenotypic variation. Under antibiotic and stress-combination conditions, fitness was driven by discrete, high-effect determinants, including known resistance genes, resulting in stronger signals and improved predictability. In contrast, non-antibiotic environments showed more polygenic and distributed architectures with weaker associations. Genome-wide analyses identified both established and previously uncharacterized genes linked with fitness across conditions. Resistance and virulence determinants exhibited clear context-dependent trade-offs, conferring fitness advantages under selection but imposing costs in non-selective environments. Consistent with this, plasmid carriage showed environment- and genotype-dependent fitness effects, with benefits under antibiotic pressure and measurable costs otherwise. Genomic variant–based models for fitness prediction achieved moderate performance (Mean Spearman correlation (ρ) = 0.36 (95% CI: 0.18–0.67) for predicted versus observed values in unseen data) across conditions, with improved accuracy under strong antibiotic selective pressures, and produced well-calibrated prediction intervals with high coverage. Despite strong population structure effect on predictions, models captured predictive gene and SNP biomarkers for fitness.

**Conclusion:** These findings highlight that bacterial fitness is an emergent property of genome–environment interactions rather than a fixed attribute of genotype. This work establishes a unified high-dimensional genotype–phenotype framework linking genomic variation to fitness across diverse conditions in a major pathogen, with broader implications for other pathogenic bacterial species.

## Introduction

Bacterial fitness emerges from genome–environment interactions, yet how genomic variation translates into phenotype across diverse environments remains a central challenge, particularly for clinically relevant traits shaped by complex and context-dependent genetic effects. This challenge is especially acute in *Klebsiella pneumoniae*, a Gram-negative opportunistic pathogen and leading cause of hospital-acquired infections worldwide. The clinical significance in *K. pneumoniae* is driven by two main threats: antimicrobial resistance and virulence. Canonical resistant strains have long been associated with high mortality. Pneumonia alone may kill up to 50% of patients even when treated, while *K. pneumoniae* infections caused by resistant strains against last resort antimicrobials carry in-hospital mortality rates as high as 60% [1]. In addition, the emergence of hypervirulent lineages, capable of causing severe invasive infections in otherwise healthy individuals, has further amplified the clinical impact of the pathogen. This clinical burden is underpinned by highly diverse population and a dynamic genome [2]. Large-scale population genomic studies of *K. pneumoniae* have uncovered intricate interactions between adaptive, resistance, and virulence determinants across diverse sequence type (ST) lineages, driven in part by the extensive mobilization of genetic elements. [3–5]. A repertoire of genetic loci has been associated with clinical significance, suggesting a complex network of genes underpinning bacterial fitness in clinical settings [2, 6]. Understanding how these genetic factors enable bacterial survival across diverse environments and drive disease requires resolving the fitness landscape of the pathogen.

The fitness landscape of *K. pneumoniae* has been mapped at the gene level across diverse strains and pathobiological contexts, revealing a dynamic, modular, and highly environment-dependent architecture. Mapping the fitness landscape has thus far employed a range of approaches, including genome-wide transposon mutagenesis (e.g., Tn-seq, TraDIS), CRISPRi-based screening, comparative fitness profiling, metabolic modelling, and trait-focused functional genomics [7–10]. These approaches have identified both core and conditionally essential genes, those required only under specific environmental or host conditions, across diverse host-relevant niches, including the lung, bloodstream, urinary tract, and gut. Consistently, capsule biosynthesis, siderophore systems, and key metabolic pathways emerge as critical for fitness [11–14]. CRISPRi has enhanced resolution by targeting essential and regulatory genes, while comparative and population genomic analyses have revealed both conserved and lineage-specific adaptations [11–14]. Computational models also predict metabolic flexibility across thousands of strains, and functional genomics has yielded fine-scale maps of virulence regulation [15]. Key insights reveal that fitness in *K. pneumoniae* is highly environment-dependent, with many genes becoming essential only under specific stresses or niche conditions, and that strain variation, functional redundancy, and adaptive trade-offs collectively shape the landscape of survival and resistance [15–18].

Despite their strengths, the abovementioned approaches have important limitations. They typically rely on narrow, static conditions that fail to reflect the diverse chemical stresses found in clinical or environmental settings, often overlooking context-dependent gene functions. These approaches also tend to assess gene essentiality under limited perturbations and rarely capture large-scale phenotypic outcomes such as growth or viability. In addition, most studies have focused on a small number of lab reference strains, neglecting the diversity of natural *K. pneumoniae* populations. Chemical genomics addresses these gaps by systematically exposing diverse strains libraries to a wide range of stress conditions to quantitatively profile their fitness responses [19, 20]. This enables the construction of high-resolution functional maps that integrate genetic and environmental complexity, revealing how genes and pathways respond to stresses. Coupling such high-throughput phenotypic screening with whole-genome sequencing provides a powerful framework for resolving the fitness landscapes of natural *K. pneumoniae* populations across diverse environments.

Here, we systematically mapped and predicted the fitness landscape of a large clinical *K. pneumoniae* collection across diverse environmental and pharmacological conditions. We profiled 1,462 clinical *K. pneumoniae* isolates for fitness under 214 diverse environmental and clinically relevant stresses, including antibiotics, chemical stressors, environmental perturbations, metabolic substrates, and combinatorial conditions. We employed genome-wide association methods and interpretable machine learning models to identify biomarkers associated with bacterial fitness across diverse strains and to link phenotypes with genomic variants. Our results show that bacterial fitness is determined by a highly polygenic, environment-dependent genetic architecture, shaped by strong genotype–environment interactions. The contribution of different genomic modalities varies across conditions, with associations highlighting the roles of resistance and virulence determinants, clinically relevant plasmids, and previously uncharacterized yet mechanistically plausible genes in shaping the fitness landscape. To our knowledge, this study provides one of the first systematic frameworks for characterizing genotype–phenotype–fitness relationships across diverse environmental conditions in clinical strains of a major pathogen, with implications extending to other bacterial pathogens.

## Methods

### Strain Collection

A library of 1,462 clinical *K. pneumoniae* complex isolates was analysed as part of this study. These isolates were predominantly recovered from a hospital network over an eight-year period through routine clinical screening as part of a multicenter genomic epidemiology study. Isolates were obtained from a range of clinical sources, including blood (63%), urine (25%), and other specimen types from different hospital wards. Isolate identification and whole-genome sequencing were performed using previously described protocols [21]. The strains were arrayed and stored in 96-well plates consisting of mutants that showed reproducible growth in LB broth. The reference strain MKP103 [22] was added as the reference control across plates. Chemical genomics conditions and genome accessions and characterizations of the isolates are provided in Supplementary Tables S1 and S2, respectively.

### Chemical genomics screen

The chemical genomics pipeline were implemented as outlined in Figure S1 and described previously [20]. The resultant data was processed using a computational/analytical pipeline (Figure S2). Strains were arrayed at high density (1,536 strains per plate) onto LB Lennox agar plates (20 g/L agar; Sigma-Aldrich) using a BM6-BC robot (S&P Robotics Inc.) and incubated at 37 °C for ∼6 hours. These library plates were then used as a source to pin the arrayed *K. pneumoniae* isolates onto condition plates containing the stressor for chemical genomics experiments.

A comprehensive panel of 214 stress conditions was designed, spanning nine categories: chemical stress, carbon source utilisation, antibiotics, environmental stress, antiseptic, stress combinations, metal stress, base (i.e. LB Lennox agar), and envelope stress (Supplementary Table S1, Figure S3). Unless otherwise specified, assays were performed on LB Lennox agar (20 g/L agar; Sigma-Aldrich), while nutrient limitation experiments were conducted on M9 minimal medium (20 g/L agar; Sigma-Aldrich). Chemical stressors were incorporated into molten agar prior to plate pouring, whereas environmental perturbations were applied during incubation. Congo red (20 µg/mL) and Coomassie Brilliant Blue (10 µg/mL) were added to all plates to facilitate colony detection. Plates were incubated at 37 °C until colonies became clearly visible (typically 6–24 h, depending on the condition). Imaging was performed under controlled illumination using an 18-megapixel Canon Rebel T3i camera mounted on the BM6-BC robotic platform (S&P Robotics Inc.) (Figure S1). Colony size was quantified using IRIS software, which detects plate boundaries, segments colonies, and measures pixel area to generate numerical outputs for downstream analysis [23].

### Chemical and environmental conditions

Conditions were selected to capture key fitness determinants of *K. pneumoniae* across clinically relevant environments and grouped into the above categories to structure experimental design and downstream analyses (Supplementary Table S1). Where *K. pneumoniae*-specific data were unavailable, conditions were informed by mechanistic studies in related Gram-negative bacteria or known physicochemical effects of the stressors and included to expand the chemical space. The condition categories included the following:

#### Antibiotics

We included a panel of 77 antibiotic conditions spanning major therapeutic classes (Figure S3). β-lactam antibiotics dominated the panel, including penicillins, cephalosporins, carbapenems, the siderophore cephalosporin cefiderocol, and combinations with β-lactamase inhibitors. Additional classes targeting core cellular processes included aminoglycosides and tetracyclines (protein synthesis), fluoroquinolones (DNA replication), and rifamycins (transcription). Membrane-targeting agents comprised polymyxins, while other classes, including amphenicols, lincosamides, glycopeptides, and lipopeptides, captured broader translational and cell envelope perturbations.

#### Chemical stress

Chemical stress conditions were grouped into five functional categories: antimicrobial/bioactive compounds, metal/inorganic stressors, organic small molecules, surfactant/membrane-active agents, and other perturbations (Figure S3). These conditions were designed to disrupt core physiological processes, including energy metabolism, membrane integrity, and efflux activity. Protonophores such as CCCP were used to collapse the proton motive force, thereby impairing aminoglycoside uptake and resistance–nodulation–division (RND) efflux systems, whereas efflux-modulating compounds such as verapamil were included to probe efflux-dependent resistance mechanisms.

#### Environmental stress

Environmental stress conditions were structured to probe physicochemical constraints across four categories: ionic/salt stress, osmotic stress, pH/buffering conditions, and physical/environmental stress (Figure S3). Ionic and osmotic perturbations included variations in salt concentration (NaCl), osmolarity (sucrose), and compatible solutes (trehalose, sorbitol, mannitol, proline), capturing adaptation to saline, desiccated, and osmotic environments. pH and buffering conditions spanned acidic to alkaline regimes (pH 5–9), including weak organic acids and bicarbonate buffering, to expose pH-dependent effects on growth. Physical and environmental stresses included oxygen limitation, temperature shifts (20 °C), and UV exposure, targeting respiration, membrane adaptation, and DNA damage responses.

#### Carbon source utilisation

Carbon source utilisation conditions were structured into four categories: carbon sources, complex nutrient, nitrogen/nucleic substrates, and stress/physicochemical conditions (Figure S3). Isolates were grown in minimal media supplemented with defined substrates, including monosaccharides, disaccharides, sugar alcohols, amino acids, and nucleosides, to resolve substrate-specific metabolic capacity.

#### Stress combinations

Stress combination conditions were structured into three categories: antibiotic–antibiotic combinations, antibiotic–environment conditions, and antibiotic–non-antibiotic combinations (Figure S3). Antibiotic–antibiotic combinations included cefepime–tazobactam and piperacillin–tazobactam, designed to restore β-lactam activity through β-lactamase inhibition; cefiderocol–ceftazidime, cefiderocol–imipenem, and cefiderocol–meropenem, which combine siderophore-mediated uptake with cell wall inhibition to overcome permeability and resistance barriers; cefiderocol–colistin and colistin–cefepime, which pair outer membrane disruption with intracellular-acting antibiotics to enhance penetration; cefiderocol–tigecycline, targeting coordinated inhibition of cell envelope synthesis and protein translation; ciprofloxacin–rifampicin, combining inhibition of DNA replication and transcription to suppress compensatory responses; and colistin–ciprofloxacin and colistin–gentamicin, which exploit membrane permeabilization to potentiate uptake of fluoroquinolones and aminoglycosides.

#### Metal stress

Metal and chelator conditions were structured within the metal/inorganic stress category to directly interrogate metal homeostasis, ion toxicity, and nutrient limitation (Figure S3). These included the metal chelator EDTA, which destabilises the outer membrane by removing Mg²⁺ and Ca²⁺ ions that bridge lipopolysaccharide, increasing permeability [24]. Iron availability was explicitly modulated using FeCl₃ to probe siderophore-dependent acquisition, a key determinant of fitness in host niches such as urine and serum [9]. Supplementation with Mg²⁺ and Zn²⁺ salts was used to perturb metal homeostasis and regulatory networks governing metallostasis, which tightly control intracellular metal pools. Toxic metal stressors included silver nitrate and sodium fluoride, which target thiol-containing enzymes, disrupt redox balance, and interfere with central metabolic processes including glycolysis and DNA integrity.

#### Envelope stress

Envelope stress conditions included non-ionic surfactants Triton X-100, Tween 20, and Tween 80, which perturb membrane integrity by inserting into lipid bilayers and altering permeability. Triton X-100 acts as a potent membrane-disrupting detergent, whereas Tween surfactants induce partial permeabilization and increase membrane transport [25].

#### Antiseptics

To capture responses to disinfectants and host-associated antimicrobial compounds, we included membrane-active antiseptics, detergents, bile salts, organic solvents [26] and cationic peptides [27]. These agents disrupt membrane integrity, capsule stability and permeability and are relevant to both clinical and environmental survival [28]. In *K. pneumoniae*, envelope integrity and efflux systems play central roles in tolerance to membrane-active compounds and host-associated stresses, contributing to persistence across both environmental and host-associated niches [29, 30].

### Fitness calculation and data analysis

For each condition, four independent replicates were imaged and analysed using Iris software to quantify colony size at each array position [23]. Colony size measurements were then processed using a standardized pipeline to derive strain-level fitness estimates while accounting for technical variation across replicate plates. The resulting data were initially processed using the ChemGAPP software, which performs multiple quality-control steps, including filtering low-quality conditions and correcting errors such as mis-pinning or mislabelling [31].

Colony coordinates were mapped to strain identities using predefined plate layouts. Colonies with zero-size measurements were temporarily excluded during normalization to avoid artefacts arising from empty, failed, or incorrectly pinned positions rather than true biological absence of growth. To correct for systematic plate effects, colony size measurements were adjusted using a linear mixed-effects model with plate included as a random effect, thereby accounting for non-independence among observations within and across plates. Adjusted colony sizes were then recentred to a common baseline, and relative fitness values were calculated by scaling each colony measurement to the median colony size of the corresponding plate. Following normalization, true zero-growth observations were reintroduced into the dataset. Replicate measurements were subsequently summarized at the strain level using the median relative fitness across plates, yielding isolate-level fitness estimates for downstream analyses.

Model-based clustering of conditions based on the relative fitness values of the collection was performed on the t-SNE embedding (two-dimensional coordinates) using Gaussian mixture models implemented in the mclust R package, with the optimal number of clusters (G = 1–20) selected based on the Bayesian Information Criterion (BIC) [32].

Reproducibility across replicates was quantified using the intraclass correlation coefficient (ICC), computed across plate-level measurements for each condition. ICC was defined as the proportion of total variance attributable to differences between subjects, calculated as 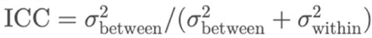, where 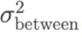 represents variance between subjects and 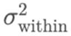 represents within-subject (residual) variance. This metric ranges from 0 to 1, with higher values indicating greater consistency among repeated measurements and lower contribution of measurement error. ICC was employed to assess the reproducibility of replicate assays and to support the use of aggregated measurements (i.e., mean values) as representative fitness measure. Hartigan’s dip test was used to assess the shape of the distribution of relative fitness across conditions and to test for deviations from unimodality using diptest library in R [33]. The test evaluates the null hypothesis of a unimodal distribution against the alternative of multimodality (e.g., bimodality), with statistical significance assessed using the p-value (p < 0.05 indicating rejection of unimodality and evidence for multiple modes).

### Genomic analysis

We used the whole genome sequencing data for the collection, which went through the microbial genomics pipelines for *de novo* assembly, annotation, variant calling as detailed previously [21]. Pangenome reconstruction was conducted with Panaroo using strict clean-mode strict and after removing invalid genes [34]. MLST, resistome and virolome chcaterization was performed using Kleborate (v2.3.2) and AMRFinderPlus [35]. Virulence and antimicrobial resistance profiles were inferred using the gene-based scoring framework implemented in Kleborate. Virulence (score 0–5) was assigned based on the presence of key loci in a hierarchical manner (yersiniabactin, *ybt*; colibactin, *clb*; aerobactin, *iuc*): 0, none detected; 1, *ybt* only; 2, *clb* alone or *ybt* + *clb*; 3, *iuc* only; 4, *iuc + ybt* without *clb*; and 5, *ybt + clb + iuc*. Resistance (score 0–3) was defined according to increasing clinical severity: 0, absence of ESBL and carbapenemase; 1, ESBL only; 2, carbapenemase without colistin resistance; and 3, carbapenemase with colistin resistance determinants. Based on these scores, isolates were classified into pathotypes: genomes with resistance score ≥1 were designated ESBL(+)/CP(+), those with virulence score ≥3 were defined as hypervirulent (hv), and genomes meeting both criteria (resistance score ≥1 and virulence score ≥3) were classified as convergent. Fitness, virulence, and resistance associations were quantified by integrating condition-specific fitness profiles with Kleborate-derived genomic scores. Single-nucleotide polymorphisms (SNPs) in the core genome were identified by aligning short-read data to the reference genome of *K. pneumoniae* MKP103 (accession PRJNA765043) using the Snippy pipeline with default parameters (www.github.com/tseemann/snippy). Annotated SNP outputs from Snippy were used for downstream analyses, including filtering for missense and stop-gain variants.

To assess the relationship between bacterial fitness and virulence and antimicrobial resistance levels, we modelled strain-level fitness values across all conditions using linear regression. For each condition, fitness was regressed against virulence score, resistance score, and their interaction term:

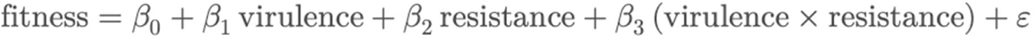

where β_0_ is the intercept, β_1_ and β_2_ represent the main fitness effects of virulence and resistance, β_3_ captures their interaction, and ε denotes the residual error.

Fitness values were derived from the normalized colony size measurements for each strain under each condition. For each condition, the regression coefficient corresponding to the virulence and resistance scores was extracted, along with its associated p-value. 95% percent confidence intervals were computed using standard Wald-based intervals. To account for multiple testing across conditions, p-values were adjusted using the Benjamini–Hochberg procedure. Associations were considered significant at an adjusted p-value < 0.05.

### Plasmid effect analysis

We identified the plasmid replicons in the short-read sequencing data with Abricate (https://github.com/tseemann/abricate) using the PlasmidFinder database [36] and the default cut-off. We retained the ten most prevalent plasmid replicons. Fitness measurements were aggregated by plasmid count to compute mean fitness values for each plasmid replicon for each condition. These values were then normalized to the mean fitness of the baseline group (isolates without plasmids) to estimate the effect of plasmid replicon number on fitness for each condition. We also computed the individual contribution of each plasmid replicon to fitness under each condition using an elastic net regression framework for the ten most frequent plasmid replicons (types), modelling fitness as a function of plasmid-associated genetic features. For each condition *j*, fitness *y^(j)^∈ℝ^n^* was modelled as a linear function of plasmid-associated features *X∈ℝ^n×p^*:

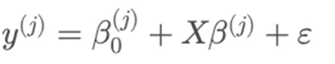

Model parameters were estimated by minimizing the elastic net penalized least-squares objective. Optimal regularization values (i.e. λ values) were selected for each condition using cross-validation (cv.glmnet) function in R, and the corresponding coefficients were extracted. These were used as plasmid replicon fitness effect for each condition.

The full coefficient matrix:

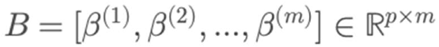

was then constructed by concatenating coefficients for *p* plasmid replicons across all *m* conditions, which allowed comparison of plasmid feature contributions across conditions.

To assess whether plasmid-associated fitness effects differed between genetic backgrounds, we performed a paired comparison for each condition and plasmid replicon type in the ST147 and ST2096 lineages, i.e. the two most frequent lineages in the collection. To this end, for each plasmid, we first obtained the plasmid replicon fitness effects in the ST2096 and ST147 subpopulations, as described above. We then computed the difference in fitness in ST2096 and ST147 lineages in each condition as Δβ = β_ST147_ − β_ST2096_. For each plasmid replicon, we quantified the overall fitness effect as the mean Δβ across all conditions. To test whether the distribution of Δβ deviated significantly from zero, we applied a two-sided Wilcoxon signed-rank test. All analyses were performed independently for each plasmid type.

### Bacterial GWAS

We employed a bacterial genome-wide association study (bGWAS) approach to identify genetic determinants associated with fitness and to perform feature engineering for the machine learning pipeline. We employed Scoary (v1.6.16) [37], which evaluates gene–trait associations using Fisher’s exact test followed by Benjamini–Hochberg correction and incorporates a phylogeny-aware pairwise comparison framework to mitigate lineage confounding. Since Scoary accepts only binary traits, we first binarized fitness values for each condition according to their median population value. Genomic predictors included accessory gene family presence–absence (defined using Panaroo), core-genome presence-absence SNPs identified with Snippy, presence-absence of unitigs (i.e., unique sequence fragments formed by merging kmers into maximal unambiguous stretches) generated by the unitig-caller pipeline (http://www.github.com/bacpop/unitig-caller) [38] and ressistance and virulence determinants identified by AMRFinderPlus pipeline. Features were first filtered based on nominal significance (p < 0.05) and false discovery rate-adjusted p-values (< 0.05). We then applied phylogeny-informed criteria, retaining features with both best and worst pairwise p-values < 0.05 for stringent (narrow) associations, or best pairwise p-values < 0.05 for broader signals.

For the accessory genomic hits, we performed additional validation by running BLAST on the consensus sequence for each gene family identified as a hit against the assemblies, applying a maximum E-value of 10^−5^, a minimum identity threshold of 90%, and a minimum query coverage per high-scoring pair (HSP) of 80%. By doing so, we confirmed that the presence–absence patterns inferred by Panaroo were not artifacts of partial contigs or gene fragments. We also used GWAS to identify pleiotropic genes, defined as accessory genes associated with multiple condition categories. To quantify this, we defined a pleiotropy score as the number of categories in which a gene showed a significant association with fitness in at least one condition within that category.

For GWAS at the SNP level, we created a table corresponding to the presence and absence of SNPs at each genomic site in the KPNIH1 reference genome. We then fed it into Scoary and filtered for stringent associations. We excluded synonymous SNPs, as they are not likely to have any functional effect. For missense SNPs, we validated the fitness effects of the significant SNPs in the LB condition by comparing them with the effects of mutations in the same genes in the single-gene deletion library assay. This allowed us to examine whether disruption of gene function by a missense mutation or transposon insertion yields similar fitness effects. To this end, we used an ordered transposon mutant library of KPNIH1 previously generated by [39]. This analysis tested whether genes harbouring significant missense variants also exhibited altered fitness when disrupted by transposon insertion. The mutant library was generated using the mini-Tn5 derivative T30 in the MKP103 background strain lacking the KPC-3 carbapenemase gene and condensed into 4,346 mutants. Mutants were screened on LB agar using the same chemical genomics pipeline described above (Figure S1). Colony images were analysed using ChemGAPP and relative fitness values were calculated as standardized Z-scores. For validation, we identified the Z-score corresponding to each gene containing a significant missense SNP and tested whether these genes were enriched among mutants with above-median fitness effects compared with random expectation. Statistical significance was assessed using a permutation framework (10,000 permutations), where the p-value was calculated as the fraction of random samples producing an equal or greater enrichment than observed.

### Population structure features and genotype-phenotype distance comparison

We used population structure (lineage) features as predictors in the machine learning models. To this end, we identified genetic clusters within the population. Pairwise Jaccard distances based on shared unitigs between isolates were computed to generate a genetic distance matrix. Genomes were then clustered using a distance-based approach implemented in the adegenet package in R [40], exploring clustering resolutions from one up to the total number of strains. The Gengraph function in the package first constructs graphs from pairwise genetic distances, where edges connect genomes below a defined distance threshold, and clusters were defined as connected components within the resulting graphs. Based on these clusterings, we obtained the population structure matrix S, where *S^ij^=k* if genomes *i* belongs to cluster *k* in the clustering with at most *j* clusters. In total 50 population structure features were identified.

Using the unitig similarity matrix, we also computed the correlation between genetic (unitig-based) and phenotypic (fitness-based) distance matrices using a Mantel test implemented in the vegan package in R, with significance assessed by permutation. The Jaccard distance matrix was used to construct a neighbour-joining tree in R using the ape package [41]. Metadata and the phylogenetic tree were visualized using ggtree [42].

### Machine learning platform

A supervised learning framework was developed to predict condition-specific fitness values from genomic features. For each condition, the response variable was defined as the corresponding relative fitness measurement, while predictor variables comprised a hierarchical combination of genomic descriptors depending on the modelling mode. These included population structure (cluster assignments), significant pangenome gene presence–absence profiles, SNPs, and unitigs identified by the bGWAS pipeline. The latter three feature sets were one-hot encoded, while a multi-encoding approach was applied to represent the population structure matrix. Four modelling modes were defined to systematically evaluate the contribution of different genomic feature layers. Mode 1 included population structure alone to capture lineage effects. Modes 2–4 incrementally incorporated pangenome, SNP, and unitig features, respectively.

To enable uncertainty-aware prediction, we evaluated a set of interval-prediction regression models, targeting the 2.5th, 50th, and 97.5th percentiles to derive 95% prediction intervals. The evaluated models included linear quantile regression as a baseline, along with the more complex ensemble methods, including gradient boosting–based quantile regression, quantile regression forests, and Natural Gradient Boosting (NGBoost) [43]. Hyperparameters were optimized using cross-validation for each model as follows: for linear quantile regression, the regularization strength (α) was tuned; for gradient boosting models, the learning rate, maximum tree depth, minimum samples per leaf, and maximum number of leaf nodes were optimized; for quantile regression forests, the number of trees (n_estimators) and maximum tree depth (max_depth) were tuned; and for NGBoost, the number of boosting iterations (n_estimators) and learning rate were optimized. The values of the hyperparameters are detailed in the GitHub page of the project.

Model training and evaluation were performed using a nested cross-validation framework. Data were partitioned into five independent splits comprising training/validation (80%) and test (20%) sets. Within each training set, hyperparameter tuning was conducted using 3-fold cross-validation, and performance metrics were averaged across folds. Final models were retrained on the full training data using optimal hyperparameters and evaluated on the corresponding held-out test sets to ensure robust generalization.

Model performance was assessed using Spearman correlation (ρ) between predicted and observed fitness values, empirical coverage of the 95% prediction intervals, and the average width of prediction intervals. To account for measurement error, correlations were adjusted as ρ / √ICC, assuming negligible error in model predictions for each condition. This yielded an upper bound on predictive performance.

### Sensitivity analysis

To assess the dependence of model performance on training set size, we performed a subsampling-based sensitivity analysis. For each condition, training subsets spanning 5–100% of the available data were generated, while the test set size was held constant (n = 50 isolates) to enable consistent evaluation. For each subset, NGBoost models were trained and tuned as described above, and predictive performance was quantified using Spearman’s rank correlation (ρ) between observed and predicted fitness values for the test and training sub-datasets. To ensure robustness, the procedure was repeated ten times with independent random subsamples at each training size. We aggregated the results for all conditions.

### Lineage-stratified evaluation

To examine model generalizability on new lineages, we performed a lineage-stratified evaluation. For each condition, training and test datasets of fixed sizes (n = 1,000 and n = 50, respectively) were constructed such that the test set comprised isolates from a single ST, while the training and validation sets included isolates from all other STs. NGBoost models were trained, tuned, and validated on the training data as described above. This procedure was repeated for the five most frequent STs, each held out in turn as an independent test set, to evaluate model performance on unseen lineages. To provide a baseline for comparison, models were also trained and evaluated on randomly shuffled (non-stratified) datasets with identical training and test sizes. Predictive performance was quantified using Spearman’s rank correlation (ρ) between observed and predicted fitness values on the test data. Differences in performance between stratified and non-stratified settings were used to assess the extent to which models capture lineage-independent signals.

### Feature important analysis

To identify predictive features in the input predictors, feature importance was quantified using SHapley Additive exPlanations (SHAP), a game-theoretic framework that assigns each feature a contribution value for individual predictions [44]. For tree-based models, SHAP values were computed using the TreeExplainer algorithm, which provides estimation of feature contributions for ensemble tree models [44]. Predictive modelling was performed using Extreme Gradient Boosting (XGBoost). Models were trained to predict condition-specific fitness values using genomic features, and hyperparameters, including the number of boosting iterations (n_estimators), learning rate, and maximum tree depth, were optimized via cross-validation. Subsampling (subsample = 0.8) and feature subsampling (colsample_bytree = 0.8) were applied to reduce overfitting and improve generalization. We ran the model on five splits of training/validation and test splits and obtained SHAP values for each split on test datasets. Mean absolute SHAP values were calculated across all samples for the features that were found important across all test data, to obtain global feature importance scores.

### Prediction of lineages from fitness profiles

Further to predicting fitness profiles from genomic variants, we examined the extent to which genotype could be inferred from fitness in an inverse predictive framework. Fitness profiles across 214 conditions were used as predictors, and the four most prevalent ST lineages were used as class labels. These lineages included ST2096, ST14, ST101, and ST147. An XGBoost multi-class classifier was trained using the multi:softprob objective, which estimates class probabilities across multiple lineages and assigns isolates to the most likely sequence type based on their fitness signatures. The model was tuned using the hyperparameters described above and evaluated across ten independent random train/test splits. Feature importance was assessed using gain-based importance and averaged across the ten runs. Gain importance reflected the contribution of each fitness condition to reducing classification error across the ensemble of decision trees.

### Data availability

We developed the ColonyExplorer portal, which contains the results from the screening assays and colony characterizations. The portal allows users to select colonies based on genome accession numbers and directly visualize growth and quantified growth features. We also provide all assay results, intermediate files, GWAS and machine learning results, along with the consensus sequences of gene families from pan-genome analysis, within the portal (https://colonyexplorer.kaust.edu.sa/ and https://github.com/gzhoubioinf/ColonyExplorer). The trained models are made available as a command-line application, allowing users to input genome assemblies and predict relative fitness under selected conditions. All R and Python codes used for modelling is also available in the project’s GitHub repository (https://github.com/Sara-Iftikhar/ChemGenomics).

## Results

### Chemical genomics screening covered diverse environmental and pharmacological conditions

To decipher bacterial fitness responses across diverse and clinically relevant growth conditions, we established and optimized a high throughput chemical genomics screening system (Figure S1, Figure 1). Using this platform, we screened a large-scale clinical collection of 1,462 *K. pneumoniae* complex isolates across 214 conditions encompassing a broad and hierarchically structured range of environmental and pharmacological perturbations (Figure S2). The assay followed a chemical genomics workflow (Figure S1), coupled with a downstream analytical framework for identifying genomic biomarkers associated with bacterial growth, which were subsequently integrated into a machine learning predictive pipeline (Figure 1). The screened conditions spanned multiple categories, with antibiotics targeting major cellular pathways representing the largest group, followed by chemical stress, carbon source utilisation, environmental stress, and stress combinations, alongside additional contributions from antiseptic, metal stress, envelope stress, and base conditions (Figure S2). Stress combinations included both antibiotic combinations and antibiotic–environmental perturbations (Figure S2).

**Figure 1.**
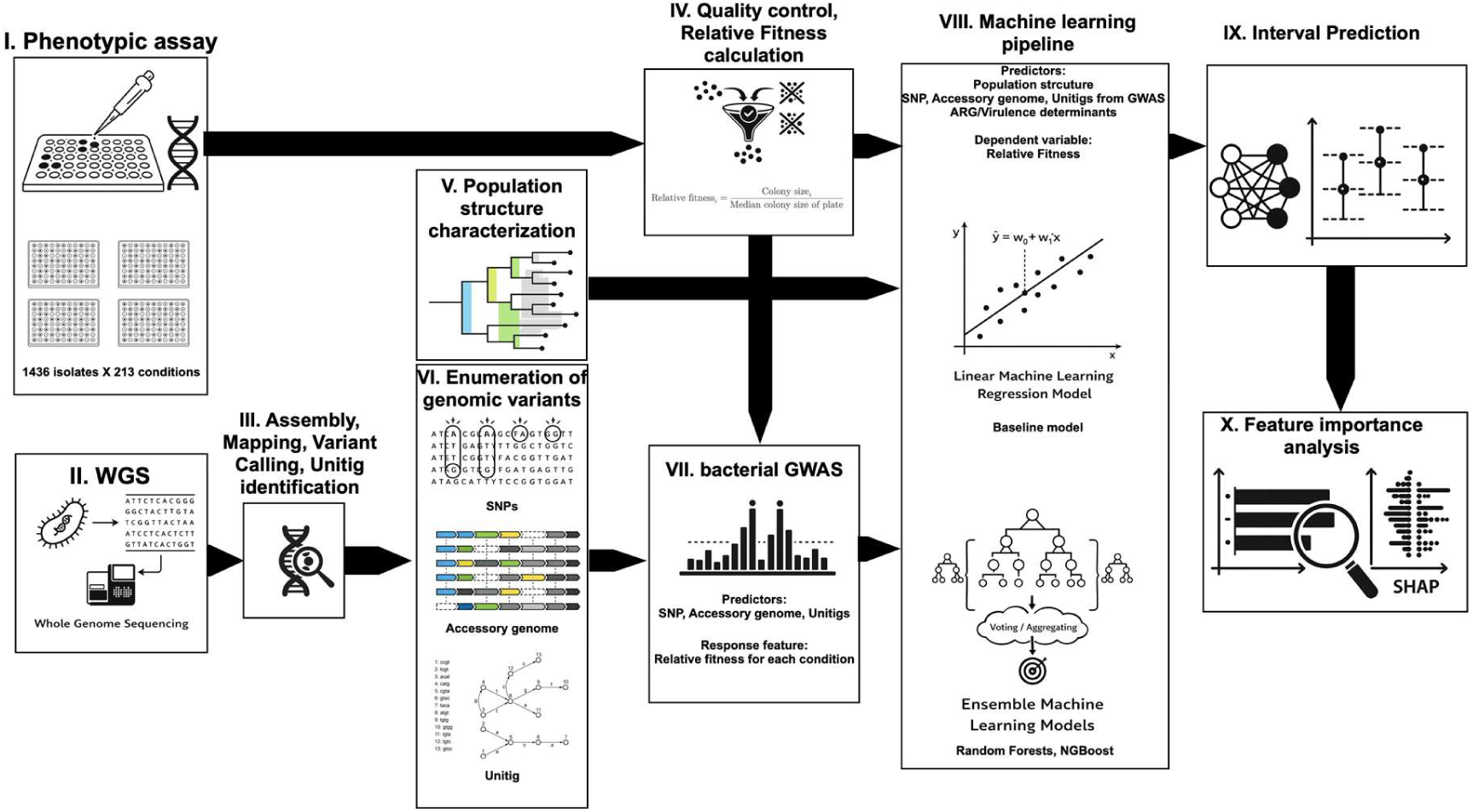
Integrated experimental and computational pipeline for large-scale genotype–phenotype mapping and fitness prediction. I–IV, Phenotypic screening and fitness estimation. A total of 1,426 bacterial isolates were screened across 214 stress conditions using high-density colony arrays. Colony growth was quantified and relative fitness was calculated as colony size normalized by the median colony size of each plate. II-III-V–VI, Genomic characterization. Whole genome sequencing data were obtained and used for downstream analyses. Population structure features were inferred from the WGS, followed by enumeration of genomic variation, including SNPs, accessory genome content, and unitigs. VII, Bacterial GWAS. Genome-wide association analyses were performed to identify significant genetic determinants associated with relative fitness across conditions, using SNPs, accessory genes, and unitigs as predictors. Significant accessory genes and SNPs were further annotated. VIII–IX, Machine learning and prediction. Predictive models were trained using genomic features (population structure, and GWAS-informed SNPs, accessory genome, and unitigs features) to predict and quantify relative fitness. Baseline and ensemble learning approaches were used, and predictive uncertainty was quantified through interval prediction. X, Feature importance analysis. Model interpretability was assessed using local importance (SHAP-based) analysis to pinpoint key genetic features contributing to fitness variation across conditions.

### Robust and reproducible fitness measurements reveal multimodal and continuous response landscapes

To ensure that this platform provides a reliable basis for fitness inference, we quantified the robustness and reproducibility of fitness measurements across conditions. To quantify strain fitness, we measured relative colony size as the pixel area of each colony. We compared this with relative opacity, which incorporates colour intensity to approximate colony thickness (area/intensity), as another measure for fitness. Both fitness proxies were strongly correlated across all conditions (Pearson’s ρ = 0.85, 95% CI: 0.78–0.89) (Figure S3A). This indicates that colony size, reflecting biomass expansion, also adequately captures variation in cell density represented by opacity. Consequently, colony size alone serves as a robust and generalizable measure of bacterial fitness. Moreover, measurements showed high reproducibility across replicates, with independent experiments exhibiting near-linear agreement across the four replicates under all conditions (Average Pearson’s ρ = 0.87, 95% CI: 0.85–0.92), as shown for two replicates in Figure S3B. Reproducibility was further supported by the intraclass correlation coefficient (ICC) values (see Methods), which remained high across most condition categories, with a mean of 0.73 (95% CI: 0.60–0.89) across conditions (Figure S4A and S4B), corresponding to moderate-to-excellent test–retest reliability [45]. Variability in ICC across condition categories suggests differences in the environmental sensitivity of fitness effects (Figure S4A and S4B). These patterns in reproducibility were paralleled by differences in the distributional structure of fitness profiles. Conditions with higher ICC values also exhibited elevated Hartigan’s dip statistics (Pearson correlation between ICC and dip statistic across conditions: 0.31, 95% CI: 0.19–0.43, p < 0.005) (Figure S4A and S4B). Higher dip values indicate stronger deviation from unimodality, reflecting more pronounced multimodal or heterogeneous fitness distributions (Figure S4A and S4B). Conditions with the highest ICC and dip values were enriched for antibiotics and antibiotic-containing stress combinations, consistent with strong and reproducible fitness responses driven by antimicrobial resistance determinants within resistant subpopulations (Figure S4A and S4B). In contrast, non-antibiotic categories with lower dip values, such as environmental and chemical stress, exhibited greater ICC variability, reflecting more continuous and context-dependent fitness landscapes.

### Clustering of fitness profiles reveals structured heterogeneity and coherent functional grouping across conditions

With robust measurements in place, we then asked whether fitness responses organize into structured patterns across environmental conditions. We therefore applied a clustering approach, which revealed structured heterogeneity across conditions, with separation into 12 distinct condition groups (Figure 2A). Antibiotic conditions were broadly distributed across eleven of the twelve clusters but showed consistent grouping in clusters 1, 11 and 12 (Supplemental table S1, Figure 2A and 2B). Clustering was biologically coherent, with β-lactams and membrane-active compounds co-grouping in cluster 2, cephalosporins dominating cluster 1, aminoglycosides and carbapenems enriched in cluster 11, and cluster 12 grouping fluoroquinolones (ciprofloxacin) with cephalosporins (cefepime), carbapenems (ertapenem) and antifolates (trimethoprim), consistent with convergent stress responses, including DNA damage and replication stress. Beyond these, clusters 8 and 2 also showed strong category-level coherence. Cluster 8 was dominated by carbon source utilisation conditions (i.e. glucose, lactose, sucrose and amino acids), indicating shared nutrient utilisation pathways. Cluster 2 contained β-lactams, tetracyclines, and several chemical stressors that produced similar bacterial fitness responses, suggesting that these conditions may activate overlapping cellular stress and adaptation pathways, as previously observed for verapamil [46]. This shared response is reflected across other clusters composed of multiple condition categories and point to common stress-response mechanisms (Figure 2A, 2B). The within- and between-condition correlation structure for fitness values also supports coherent fitness responses within condition categories, with a tendency for within-category correlations to exceed between-category correlations (Figure 2C). Antibiotics cluster with stress combinations but remain relatively structured, consistent with their specific modes of action [47], whereas environmental conditions exhibit more diffuse patterns, reflecting shared and overlapping stress-response mechanisms (Figure 2C).

**Figure 2.**
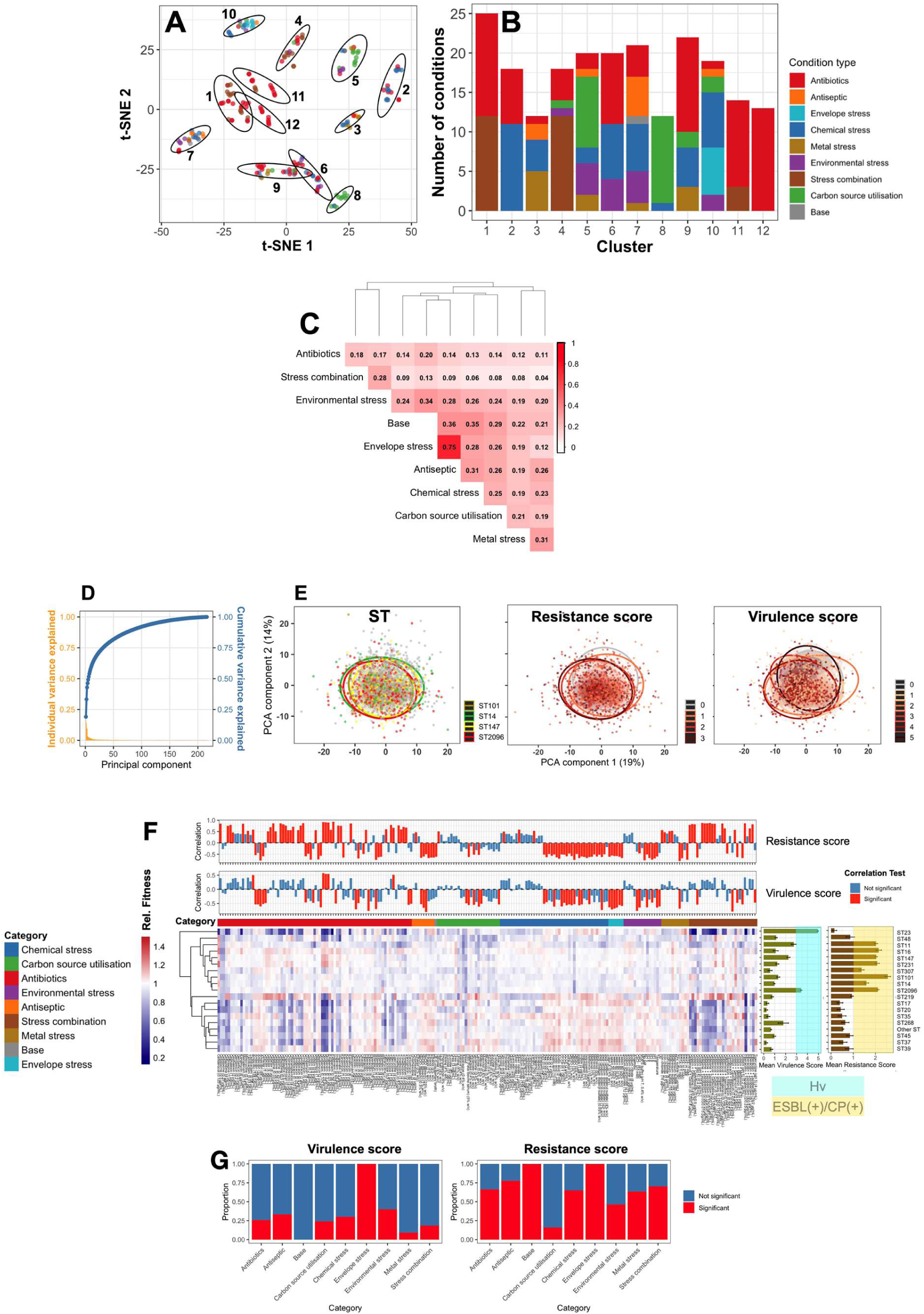
Clustering of fitness profiles across conditions. A) t-SNE projection of relative fitness profiles across all conditions, coloured by condition category type. Model-based clustering identified 12 distinct clusters. Ellipses represent the Gaussian components inferred by the model, capturing the centre and covariance structure of each cluster. The optimal model corresponded to the EEV covariance structure (ellipsoidal, equal volume and shape), selected based on Bayesian Information Criterion (BIC). **B)** Composition of each cluster by condition category type. **C)** Category-level correlation structure of fitness profiles across condition types. The heatmap shows the mean pairwise correlation of fitness profiles within and between condition categories, with hierarchical clustering highlighting similarity relationships among condition types. **D)** Principal component analysis (PCA) variance decomposition of isolate fitness profiles measured across 214 experimental conditions. Orange bars indicate the variance explained by individual principal components, while the blue curve represents cumulative variance explained. **E)** PCA projection of isolate phenotypic/fitness profiles based on the first two principal components. The left panel shows isolates colored according to major sequence types (STs). The middle and right panels show the same phenotypic space colored by resistance and virulence scores, respectively. Scores were derived using Kleborate based on the presence of key resistance determinants (carbapenemase (CP), colistin resistance, ESBL genes/mutations) and hypervirulence-associated siderophore loci. Ellipses represent the phenotypic occupancy regions of each group. **F)** Heatmap showing the relative fitness landscape across conditions for the most frequent 18 sequence types (STs) clones, with values representing mean fitness for the isolates in the clone across conditions. Columns that correspond to individual conditions grouped by category (top annotation), and rows represent STs clustered based on similarity in fitness profiles. Colour intensity indicates average relative fitness (blue, reduced; red, increased) for isolates in each ST. Bar plots on the right summarize the mean virulence and resistance scores for each ST, with error bars indicating standard deviation. The upper panels show condition-wise correlations between fitness and resistance (top) or virulence (bottom) scores, with significant associations highlighted in red. These associations between ST mean resistance and virulence scores and their corresponding mean fitness across conditions were assessed using Pearson correlation, with significance defined at pvalue < 0.05. **G)** The proportion of conditions showing significant versus non-significant correlations with virulence (left) and resistance (right) scores across condition categories.

### Broad phenotypic heterogeneity is constrained by lineage-specific MDR and hypervirulence determinants

To test whether structured phenotypic patterns reflect underlying genomic composition, we integrated fitness values with WGS data. At the population level, genetic and phenotypic distance matrices were correlated (Mantel test, r = 0.138, p-value = 0.001), showing a modest but significant alignment between genomic background and fitness profiles. The phenotypic diversity across the collection was also moderately captured by the first two principal components (33% of total variance explained), pointing to the multidimensional structure of phenotypic variation (Figure 2D). However, this structure was less strongly reflected by lineage composition, as major lineages exhibited substantial overlap in phenotypic space. In contrast, multidrug-resistant (MDR) and hypervirulence (hv) risk groups, defined using scores based on the presence of key genomic determinants of MDR (including colistin resistance, ESBL, and carbapenemase [CP] genes or mutations) and hypervirulence (siderophore loci), displayed greater phenotypic separation (Figure 2E). Analysis of major clones (sequence types [STs]; ≥1% frequency) further revealed heterogeneity in growth responses across conditions. Although ST lineages tended to cluster according to genetic relatedness, they clustered more strongly according to shared phenotypic MDR and hypervirulence risk levels (Figure 2F). The common STs represent clinically relevant lineages commonly associated with hospital- and community-acquired infections [3, 5]. ST23, a canonical hypervirulent (hv) lineage harbouring key hypervirulence siderophore loci [4, 47], formed a distinct branch, where it showed slight fitness advantage under environmental and chemical stress conditions but a lower fitness under antibiotic conditions (Figure 3A). Classical MDR lineages (ST11, ST14, ST16, ST101, ST147, ST231, ST307) [48], commonly carrying ESBL or CP determinants, exhibited a coherent phenotypic cluster (Figure 2F). ST2096, a recently emerged hv-MDR clade derived from ST14 and co-harbouring hv (i.e. aerobactin) and CP (i.e. OXA-48-like) determinants, clustered with the ancestral clone within the MDR group [21, 49]. Other non-MDR clones formed a distinct cluster (Figure 2F), suggesting that resistance and virulence background strongly shapes fitness architecture across lineages.

**Figure 3.**
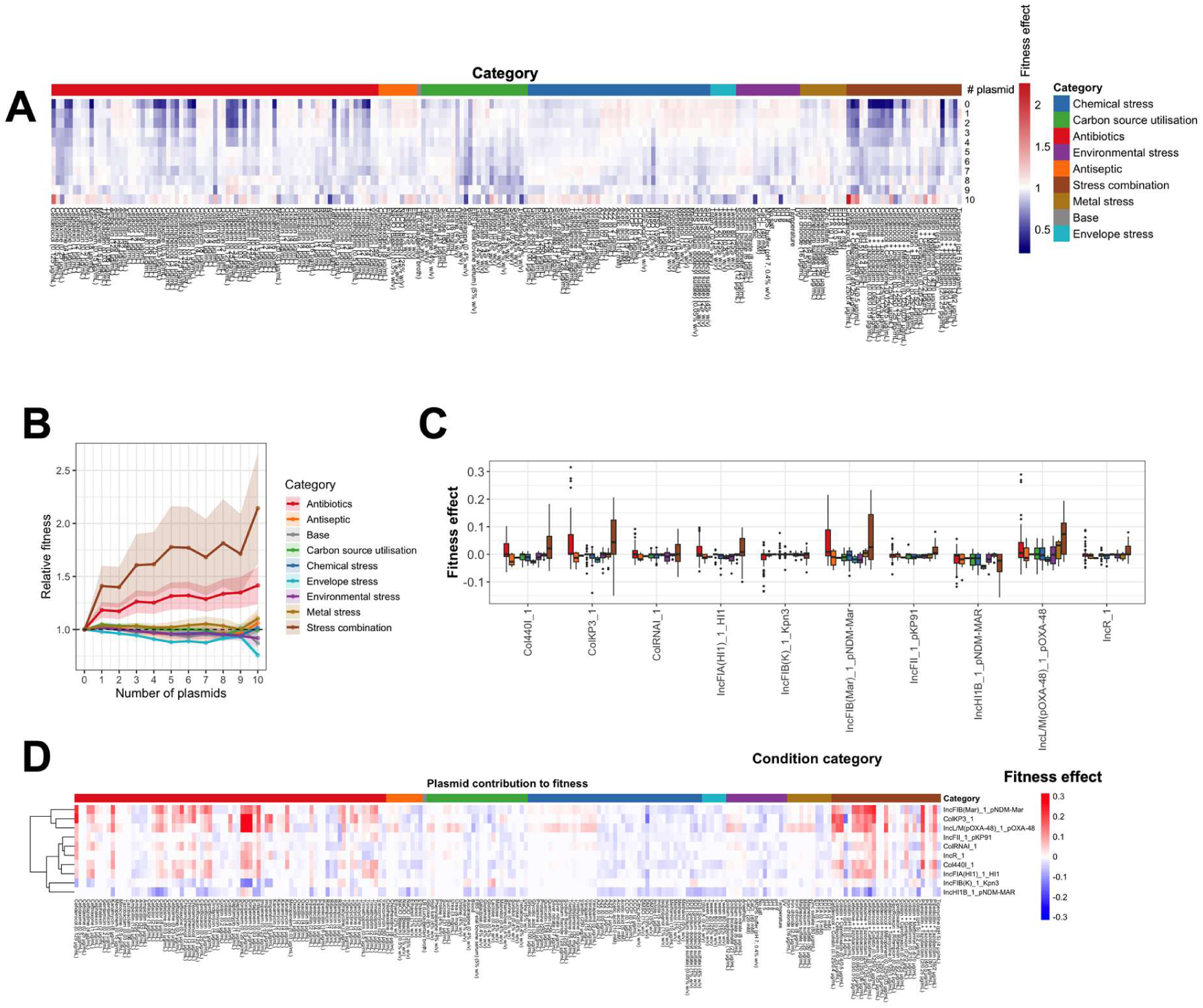
Fitness effects associated with plasmid burden across environmental conditions. **A)** Heatmap of condition-specific fitness effects stratified by plasmid replicon count for the top ten most common plasmid replicon types in the collection. Each column represents an individual condition, grouped by category (top annotation), and each row corresponds to the number of plasmid replicons (0–10). Colour intensity reflects relative fitness (blue, reduced fitness; red, increased fitness). **B)** Relationship between the number of detected plasmid replicons and relative fitness across condition categories. Fitness is normalized to strains without plasmids (reference = 1). Lines represent mean relative fitness within each category, and shaded ribbons indicate 95% confidence intervals. **C)** Aggregated distribution of fitness effect coefficients associated with plasmid replicons, aggregated across all conditions. Boxplots summarize the distribution of coefficients for a given plasmid replicon across all conditions. **D)** Heatmap of the estimated plasmid-specific fitness effects across individual conditions. Rows correspond to plasmid replicons and columns to conditions grouped by category (top annotation). Colour intensity reflects the magnitude and direction of the regression coefficient (red, positive; blue, negative), with positive and negative values indicating fitness-enhancing or fitness-reducing effects, respectively.

### Context-dependent fitness trade-offs between resistance and virulence

The MDR resistance and hypervirulence risk scores appeared to interact to shape fitness across environments. Integrating phenotypic profiles with these scores revealed distinct fitness patterns across lineages (Figure 2F, 2G). As expected, relative fitness of isolates under antibiotic and antibiotic-containing stress conditions were positively associated with resistance score. In contrast, under non-antibiotic conditions, higher resistance scores were generally linked to reduced fitness. This negative association was observed in 74% of non-antibiotic conditions, with 68% reaching statistical significance (Figure 2F). Virulence scores showed a similar pattern, with higher virulence associated with reduced fitness under environmental conditions but modest positive associations under antibiotic exposure (Figure 2F, 2G). These parallel patterns for resistance and virulence may merely reflect co-occurrence of their underlying determinants within shared genetic backgrounds [21]. To disentangle the effects of resistance and virulence, we therefore modelled fitness as a function of resistance and virulence scores, including their interaction (see Methods). Virulence still retained significant associations in 24 conditions, with most positive effects observed under antibiotic exposure (8/9 conditions) (Figure S5A), indicating a partially independent contribution of virulence-associated genes to higher fitness in these conditions. Virulence also exhibited resistance-independent fitness costs in 15 non-antibiotic conditions (Figure S5A). Interaction effects between virulence and resistance were predominantly negative (22/23 conditions) and largely confined to antibiotic conditions (Figure S5B), indicating antagonistic interactions in which their combined presence reduces fitness. Together, these findings suggest that virulence imposes condition-dependent fitness trade-offs, with effects that can be either beneficial or costly independently of resistance.

### Plasmid fitness effect depends on environment and bacterial host background

Given that the dissemination of resistance and hypervirulence determinants in *K. pneumoniae* is largely driven by plasmid-mediated horizontal gene transfer, we dissected how plasmid carriage shapes fitness across diverse environmental conditions. Stratifying fitness by plasmid burden, approximated by the number of plasmid replicons, revealed substantial heterogeneity across condition categories (Figure 3A). Plasmid-associated fitness effects were broadly monotonic but diverged sharply between antibiotic and non-antibiotic environments (Figure 3B). Under antibiotic conditions, increasing plasmid number conferred substantial fitness gains (53–203%), with the strongest effects observed under antibiotic combinations, relative to plasmid-free isolates (Figure 3A, 3B). In contrast, under less selective conditions, plasmid accumulation was linked with modest fitness costs (up to 23%), particularly under environmental and envelope stresses (Figure 3B), whereas plasmid carriage had minimal effects under chemical stresses. This highlights that the fitness consequences of plasmid carriage are environment dependent and modulated by interactions between plasmid-encoded functions and external stressors. Consistent with this, estimating the fitness effects of the ten most common plasmid replicon types in the collection within a unified modelling framework (see Methods) confirmed that these effects were context-dependent, shifting from beneficial to deleterious or neutral across conditions (Figure 3C and 3D). Replicons exhibiting the largest shifts between antibiotic and non-antibiotic conditions were predominantly associated with clinically significant antimicrobial resistance genes, particularly CP and ESBL determinants, often carried on shared plasmid backbones. This is consistent with previous genomic analyses of this collection, in which full plasmid sequences were retrieved through long-read sequencing, demonstrating that specific replicon types preferentially carry resistance determinants [21, 49]. In particular, IncFIB(Mar)_1_pNDM-Mar plasmids, occasionally harbouring ESBL genes, CP *bla*_NDM1_ and *bla*_NDM5_ genes as well as hv aerobactin loci, and pOXA-48_K8 (IncL/M(pOXA-48)_1_pOXA-48) plasmids, mostly carrying CP *bla*_OXA-48_ and occasionally ESBL genes, exhibited the strongest fitness enhancement under antibiotic conditions, with modest increases also observed under metal stress (Figure 3C and 3D). Other resistance-associated replicons, including ColKP3 (consistently carrying CP *bla*_OXA-232_) and IncFIA(HI1)_1_HI1 (frequently associated with CP *bla*_NDM1_), showed similar but weaker fitness gains under antibiotic exposure or combinations (Figure 3C and 3D). Among these plasmids, fitness benefits were most pronounced under carbapenem and cephalosporin conditions, whereas fitness costs were observed under non-antibiotic stresses, particularly chemical stressors (Figure 3C, 3D). Other plasmids showed weaker effects: IncR_1 and IncFIB(Mar)_1_pNDM-Mar plasmids, which were reported to occasionally carry ESBLs, exhibited only modest fitness increases under cephalosporin conditions. Plasmids lacking known resistance genes, such as Col440I_1 and ColRNAI_1, also showed similar patterns, with their fitness effects under antibiotic conditions likely reflecting hitchhiking through co-occurrence with resistance determinants in high-risk clones (Figure 3C, 3D). Beyond condition dependence, plasmid-associated fitness effects also varied across genetic backgrounds. Five of the ten most common plasmid types, primarily associated with resistance and virulence determinants (Col440I_1, IncFIA(HI1)_1_HI1, IncHI1B_1_pNDM-MAR, pOXA-48_K8, and IncR_1), showed significant differences in fitness effect distributions between ST147 and ST2096 isolates (Wilcoxon test, p < 0.05), indicating that the fitness consequences of plasmid carriage are strongly modulated by host genomic background.

### Resistance and virulence determinants drive context-dependent fitness trade-offs

To dissect the genetic architecture of fitness variation, we performed genome-wide association analyses using genomic variants as predictors of relative fitness across all conditions. We identified 4,143 accessory genes and 12,374 missense SNPs significantly associated with fitness in at least one condition (Figure S6A, S6B). Associations were unevenly distributed, with antibiotic and antibiotic combination conditions showing the highest number of significant hits, a pattern consistent across both pangenome- and SNP-based analyses (Figure S6A, S6B). In contrast, 23% and 26% of conditions showed no significant associations with accessory genes and SNPs, respectively, with such cases more frequent in non-antibiotic categories, including environmental stress, chemical stress, and carbon source utilisation (Figure S6B). This heterogeneity indicates that the genetic architecture underlying fitness responses varies across conditions, with stronger selective pressures, such as antibiotic exposure, generating more detectable genetic associations across lineages, consistent with bGWAS studies reporting elevated heritability and stronger genetic signals for resistance traits under selection [50–52]. Associations across accessory-gene functional classes revealed clear fitness trade-offs between environments, highlighting the context dependence of fitness architecture (Figure S7A). Significant associations were enriched for antimicrobial resistance, stress-response, and mobile genetic element genes, alongside many uncharacterized loci. The direction of association varied across environments, with the same functional classes contributing positively or negatively depending on context. Under antibiotic and antibiotic combination conditions, resistance determinants were consistently positively associated with fitness (OR > 1; Figure S7A). In contrast, across non-antibiotic conditions, including antiseptic, environmental, chemical, and envelope stresses (58/78 conditions), resistance genes were negatively associated with fitness, suggesting fitness trade-offs (OR < 1; Figure S7A). Virulence-associated genes were also consistently linked to reduced fitness across non-antibiotic conditions (OR < 1; Figure S7A). In line with this, we specifically examined significant antimicrobial resistance and virulence genes/mutations identified as hits by bGWAS and found similarly strong condition-specific fitness effects. Key MDR determinants, including ESBL *bla*_CTX-M-15_ and CP *bla*_OXA-48_ genes, conferred fitness advantages under third-generation cephalosporin and carbapenem conditions, respectively, (Figure S7B) but were negatively associated with fitness across most non-antibiotic conditions. Similarly, hypervirulence-associated siderophore genes, including aerobactin (*iucABC*) and yersiniabactin (*ybt*), showed consistent fitness costs across conditions. Aerobactin genes (*iucABC*) were also found among the most pleiotropic accessory genes (pleiotropy score = 9), showing associations with reduced fitness in at least one condition across all nine condition categories (Figure S8; Table S3). Other highly pleiotropic genes spanned diverse functional classes, including metabolic and transport systems, transcriptional regulators, and cell envelope and stress response functions, suggesting that fitness variation is associated with multiple core cellular processes across conditions (Figure S8; Table S3). Overall, these findings demonstrate that the genetic architecture of fitness in *K. pneumoniae* is polygenic and context-dependent, with resistance and virulence determinants conferring environment-specific benefits that are offset by fitness costs outside selective conditions.

### Condition-specific bGWAS uncovers causative and mechanistically plausible genes

We next assessed the utility of bGWAS in identifying functionally relevant genes by examining selected conditions, including an antibiotic, a carbon utilisation, and baseline LB growth conditions. The resulting hit lists recovered multiple known functional and antimicrobial resistance genes, supporting the biological validity of the approach. Under antibiotic pressure, fewer but clearer signals were observed. For example, under cefiderocol exposure, a siderophore cephalosporin, seven accessory genes were identified, including two well-characterised resistance determinants (ESBL *bla*_CTX-M-15_ and CP *bla*_NDM-5_) and five mobile genetic elements/hypothetical linked loci. The resistance genes showed strong associations with increased fitness (odds ratios of 5.61 for *bla*_CTX-M-15_ and 8.21 for *bla*_NDM-5_) (Figure 4A). Under nutrient-selective conditions, known functional genes were also identifiable but with weaker associations. In lactose conditions, *lacY* (lactose permease) and *lacI* (transcriptional repressor) were among the top hits (odds ratio = 4.83), consistent with their roles in facilitating carbohydrate uptake under lactose selection. These genes are dispensable in most environments but required under lactose-specific conditions. All well-characterised significant genes identified showed broad phylogenetic distribution across independent lineages (Figure 4B). In contrast to the selective conditions, other conditions exhibited weaker, more polygenic architectures, where bGWAS highlighted mechanistically plausible candidate genes rather than strong single-locus signals. For example, under the base LB condition, 79 genes showed significant associations (p-values from GWAS < 0.05). Excluding mobile genetic elements, several candidate genes were identified where absence was linked to higher fitness (Figure 4A, 4B). Among the hits, absence of phosphotransferase system (PTS) components and the maltoporin *lamB* genes was associated with increased fitness (Figure 4A). Although direct evidence in *K. pneumoniae* is lacking, this association is mechanistically plausible, as these systems mediate carbohydrate uptake and are typically induced under nutrient limitation. Under nutrient-rich conditions, their expression may impose metabolic and regulatory costs, consistent with observations that perturbation of the PTS can enhance metabolic efficiency [53, 54]. In addition to accessory genes, we identified 83 missense SNPs in core genes associated with increased growth in LB. Fitness values for the deletion of these genes, when disrupted in the *K. pneumoniae* KPNIH1 transposon mutant library, under LB condition were significantly enriched in the upper tail of the distribution (permutation test, p < 0.05). This concordance shows that missense variants identified by bGWAS are also positively associated with growth effects, captured in forward genetic screens. Genes harbouring missense variants under LB condition were enriched in core functional categories, including metabolism, nutrient acquisition, cell envelope biogenesis, transport systems, transcriptional regulation, and stress responses. Some of these variants were identified in mechanistically plausible genes such as *murB, alr*, and *murP*, implicating cell wall biosynthesis and recycling processes in growth variation under rich media conditions. These genes are associated with key steps in peptidoglycan metabolism, including precursor synthesis (*murB*), D-alanine production (*alr*), and cell wall recycling and transport (*murP*) (Figure 4A, 4B) [55]. The analyses highlight the contribution of the accessory genome to phenotypic diversity and identify candidate genes and SNPs with plausible functional relevance. To support interpretation, we provide searchable results through the ColonyExplorer portal (www.colonyexplorer.kaust.edu.sa), enabling direct inspection of colony images and annotations for isolates carrying GWAS hits (Figure 4C).

**Figure 4.**
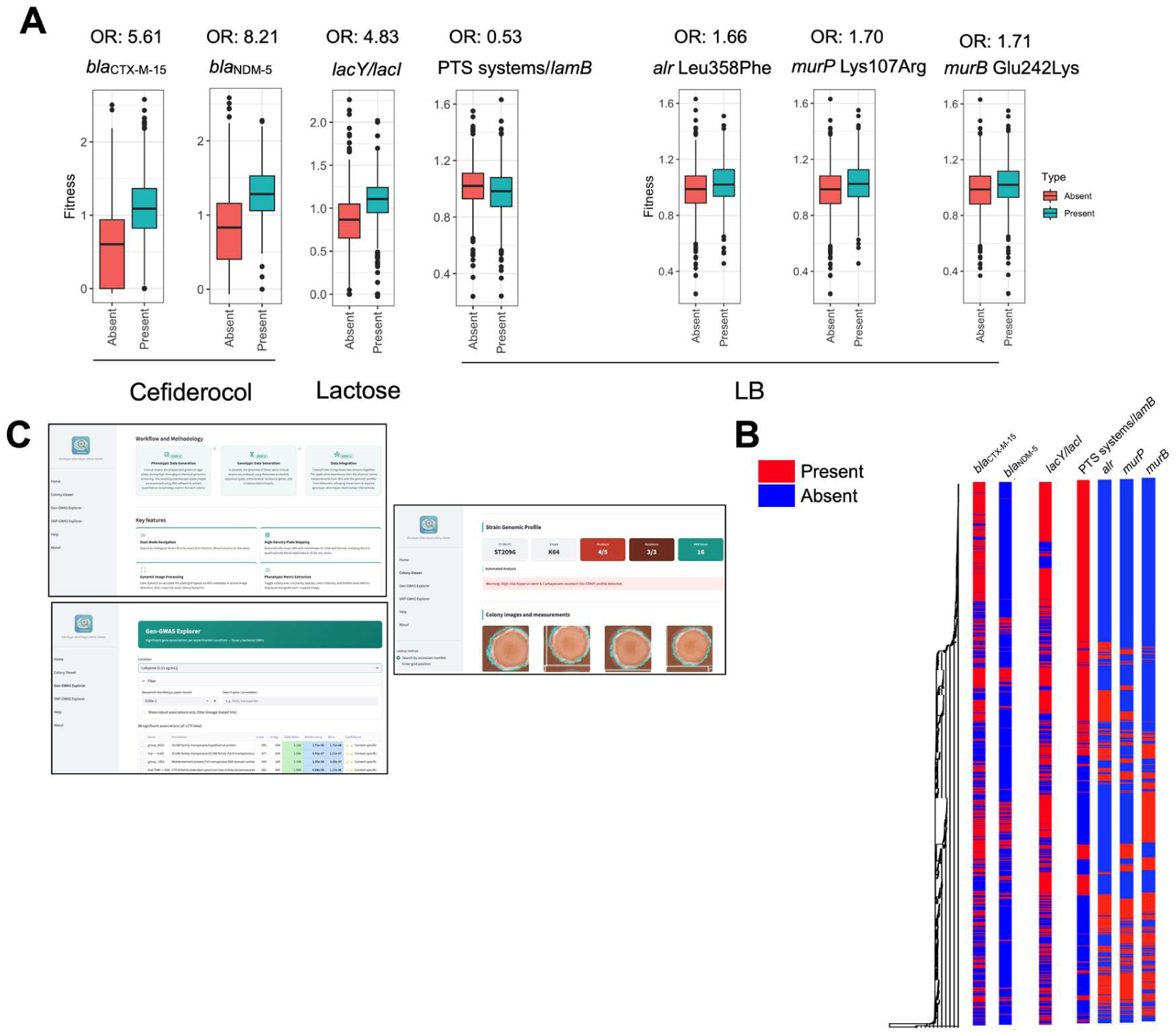
Examples of hits from bGWAS analysis from the accessory genome and SNPs levels. **A)** Distribution of fitness effects for top annotated genes across three representative conditions: antibiotic (cefiderocol), non-antibiotic base (LB), and carbon utilisation (lactose). Boxplots compare fitness between isolates lacking (red) or carrying (blue) the gene. OR denotes odds ratio. **B)** Phylogenetic distribution of selected GWAS hits across the population. The tree represents a neighbour-joining reconstruction based on the unitig distance matrix, with genes/SNPs presence (red) and absence (blue) mapped onto the phylogeny. **C)** ColonyExplorer application, with key functionalities, including visualization of colony phenotypes using strain IDs and integration of GWAS results from gene- and SNP-based analyses, enabling direct inspection of colonies harbouring GWAS hits.

### Moderate fitness prediction from genomic data, with improved performance under antibiotic and strong stress selection

We next employed bGWAS-derived genetic signals, together with population structure features, to predict relative fitness from genomic data. Model performance was assessed as the correlation between predicted and observed values, with uncertainty quantified using prediction intervals defined by their coverage probability and average width. Coverage probability reflects how often observed values fall within the predicted range, while average width captures the precision of these predictions. Models incorporated genetic information across multiple scales, from population structure to increasingly genomic features, including the pangenome, SNPs, and unitigs. We tested models of increasing complexity, from linear quantile regression to quantile random forests (QRF), gradient boosting–based quantile models (QBoost), and NGBoost (see Methods). The latter three are ensemble-based approaches, included based on prior studies demonstrating robust performance in predicting phenotypic traits from microbial genomic data [56, 57]. The models, illustrated in Figure 5A for an example fitness prediction under carbapenem (doripenem) treatment, produced point estimates that showed systematic but imperfect agreement with observed values, while accompanying prediction intervals captured the broader range within which true values are expected to lie. Across models and hyperparameter settings, incorporating richer genomic features led to incremental improvements in predictive performance, increasing the correlation between predicted and observed values by up to 20% relative to models based on population structure alone. This was accompanied by higher coverage and narrower prediction intervals when all genomic predictors were included, supporting the added value of lineage-independent bGWAS signals (Figure 5B). While overall predictive performance was broadly comparable across models, ensemble approaches (QBoost and QRF) achieved higher coverage, indicating more reliable uncertainty estimates, whereas NGBoost consistently produced narrower intervals, reflecting greater precision (Figure 5B). Selecting the best-performing model per condition based on predictive correlation revealed condition-specific performance, with NGBoost and QBoost most frequently outperforming other methods (Figure S9). Overall, best models across input predictor datasets achieved moderate predictive performance (mean Spearman correlation, ρ = 0.36; 95% CI: 0.18–0.67; range: 0.10–0.76), with high 95% coverage (mean: 0.90; 95% CI: 0.83–0.96; range: 0.80–0.98), showing that 95% prediction intervals captured the true values in ∼90% of cases across conditions (Figure 5C). Prediction intervals were moderately wide when normalized by the standard deviation (mean: 3.2; 95% CI: 2.27–4.18; range: 1.87–6.0), reflecting well-calibrated uncertainty with robust coverage across conditions (Figure 5C). Performance, calibration, and precision were generally stronger under antibiotic and stress-combination conditions, consistent with stronger predictive signals under selective pressure (Figure 5C). The moderate predictive power of genotypic data motivated us to further assess whether high-dimensional phenotypic data could be used to predict genotypic variation in a reverse prediction framework. We therefore examined the extent to which genotypic variation is predictable from phenotypic profiles by using fitness profiles to classify isolates into the four common lineages in the collection: ST2096, ST14, ST147, and ST101. Predictions on held-out datasets showed strong concordance between predicted and true lineage labels, indicating that fitness profiles captured lineage-specific discriminatory signals among dominant clonal groups (Figure 5D). Misclassifications, however, also occurred between specific lineage pairs, reflecting the shared phenotypic characteristics. The overall discrimination of the classifiers remained moderate (mean ROC–AUC = 0.74; 95% CI: 0.72–0.77 across ten random train/test splits), indicating that phenotypic profiles contain modest but detectable lineage-associated information.

**Figure 5.**
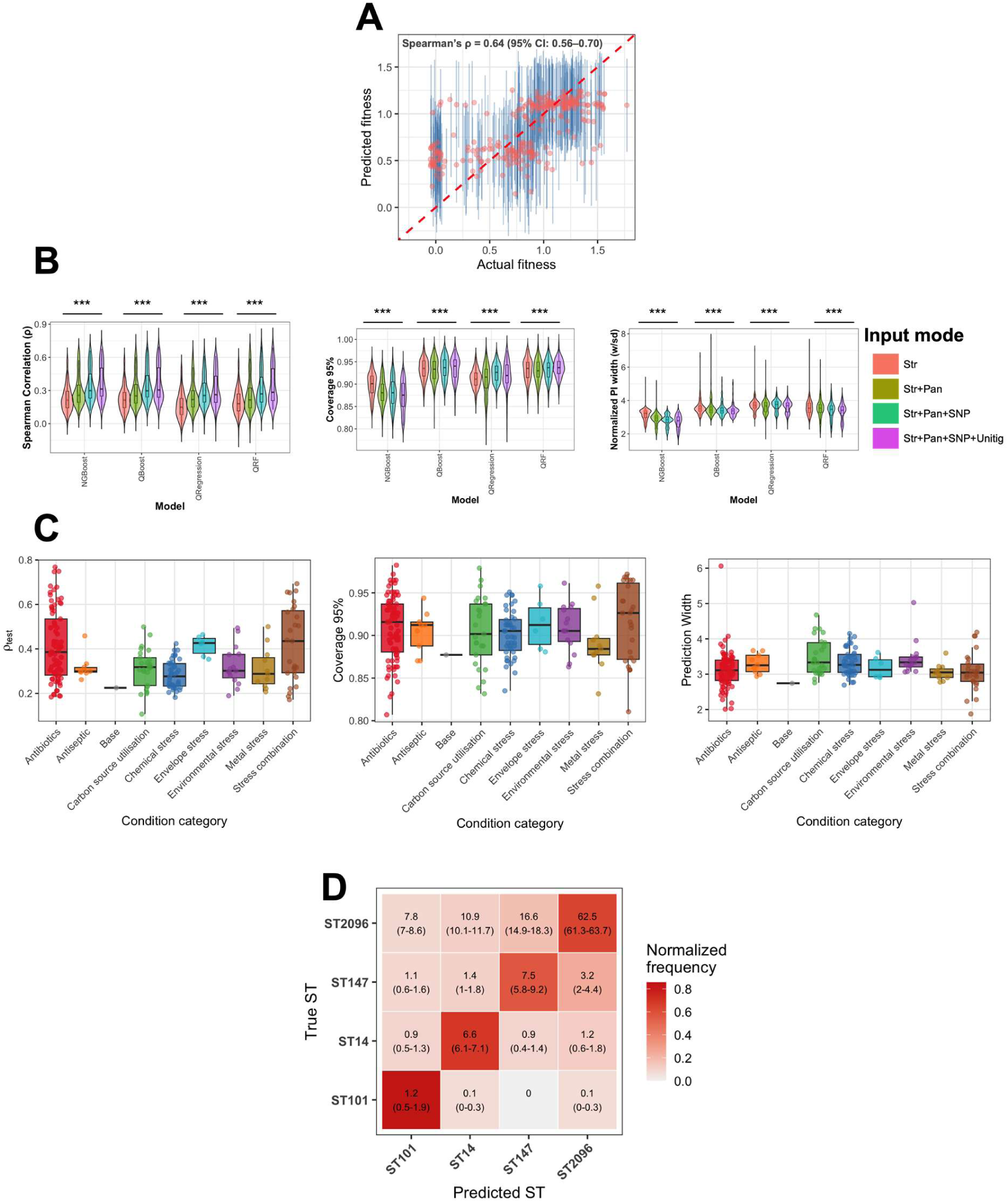
Predictive performance and uncertainty calibration across models and feature sets. **A)** Example interval fitness prediction for high concentration of doripenem, showing predicted versus observed fitness values in the held-out test dataset. Each point represents an individual strain, with vertical lines indicating the corresponding 95% prediction intervals. The dashed red line denotes the identity line. **B)** Aggregated performance metrics across all conditions, models and hyperparameters. Left, distribution of Spearman correlation coefficients (ρ) on the test datasets, summarizing predictive accuracy. Middle, empirical coverage of the nominal 95% prediction intervals, indicating calibration of uncertainty estimates. Right, normalized prediction interval width (scaled by the standard deviation of observed fitness), reflecting prediction precision. Results are stratified by modelling approach (NGBoost, quantile boosting, quantile regression, and quantile random forest) and feature sets of increasing complexity (from strain-level features to inclusion of pangenome, SNPs and unitigs). The *** signs denote statistical significance at p < 0.001, based on Wilcoxon rank-sum tests. **C)** Distribution of predictive performance metrics across condition categories for the best-performing models trained on genomic data on the test dataset. Left, Spearman correlation (ρ) between predicted and observed fitness values in the held-out test datasets. Middle, empirical coverage of nominal 95% prediction intervals, indicating calibration accuracy. Right, normalized prediction interval width (scaled by the standard deviation of observed fitness). Spearman correlation (ρ) values are normalized by the measurement errors (ICC values). Each point represents a condition, and boxplots summarize distributions within each category. **D)** Confusion matrix of lineage prediction using an XGBoost classifier trained on phenotypic profiles across 214 experimental conditions. The classifier model was trained to predict major STs from fitness measurements and evaluated across ten independent random train–test splits. Values within each cell indicate the mean number of classifications across splits, with 95% confidence intervals shown in parentheses. Cell colours represent row-normalized frequencies.

### Disentangling lineage effects in prediction reveals generalizable signals

To assess the robustness and generalizability of predictive signals, we performed complementary analyses examining sensitivity to training sample size and the impact of lineage structure on model performance. Interpolation of model performance across subsets of the training data for each condition showed a steady improvement in predictive accuracy as the training set size increased, along with improved generalizability, reflected by a reduced gap between training and test performance. However, performance exhibited diminishing returns beyond 50% of the total training data, with only a marginal (5%) improvement when increasing from 50% to 100%, suggesting that the current dataset is sufficiently large to capture most of the predictive signal with similar genomic diversity (Figure S10A). Strong population or sampling structure can confound causal inference in bacterial genomics and this pose a challenge for the application of predictive modelling [58]. To assess this effect in our predictions, we compared predictive performance under stratified and non-stratified settings. In the stratified settings, for each condition, one lineage (ST clone) was left out as unseen data, and the model was trained on the remaining STs to evaluate performance on novel lineages. Across five test clones and all conditions, the model achieved significant predictive performance in 84% of cases (i.e. correlation between actual and predicted data was significantly above zero), indicating that it captures signal that generalizes beyond closely related strains (Figure S10B). However, across most conditions, non-stratified models, where closely related strains were present in both training and test sets, showed higher apparent performance than stratified models of equivalent training size. This improvement under antimicrobial conditions likely reflects the concentration of resistance determinants within specific clones, allowing models to leverage similarities between related strains. In contrast, this effect was much weaker in non-antibiotic conditions, where the signals driving predictions are more spread out and not confined to specific clones (Figure S10B).

### Predictive models pinpoint functionally relevant genetic determinants of fitness

Finally, to assess the utility of predictive models in identifying predictive genetic biomarkers, we examined feature importance within predictive models. We dissected feature contributions to fitness prediction across conditions using SHAP values, which quantify the contribution of each feature to model predictions, for models incorporating population structure alongside GWAS-significant accessory genome features and core SNPs (Figure S11A). While accessory genome and SNP features were included as predictive inputs in most conditions (87% and 84%, respectively), population structure features, i.e. features capturing lineage effects across multiple genetic resolutions, accounted for 56% of top features (defined as those contributing 90% of total importance) across conditions and represented the most predictive feature in 148 conditions (Figure S11A). Despite this, accessory genome and SNP features were also consistently retained among the top predictors (39% and 5% of top features, and the most predictive feature in 64 and 2 conditions, respectively), indicating that models capture genetic variation that extends beyond lineage structure and is shared across independent lineages (Figure S11A, S11B). Known resistance genes emerged as the most frequent top predictors, including *bla*_CTX-M-15_ (seven conditions) and *bla*_OXA-232_ (24 conditions). This shows their strong, condition-specific associations under antibiotic pressure. In contrast, for non-antibiotic conditions, a broad and largely non-overlapping set of accessory genes and SNPs (n = 415) were identified as top predictors. Examination of the reverse prediction framework, in which genotypic lineages were treated as response variables and phenotypic profiles as predictors, identified conditions associated with strong selective pressure as the most informative features for lineage discrimination (Figure S11C). Carbon utilization traits, antibiotic responses, and combinations of stress conditions consistently ranked among the strongest predictors of lineage identity (17 out of 20 top predictor conditions), suggesting that the genotype–phenotype links are mostly shaped by selective environments.

## Discussion

In this study, we present the first large-scale chemical genomics atlas of *K. pneumoniae*, profiling over 1,462 clinical isolates across 214 diverse conditions, including antibiotics, environmental perturbations, chemical and combinatorial stresses. By integrating high-throughput phenotyping, whole-genome sequencing, GWAS, and machine learning in a unified framework, we demonstrated that bacterial fitness is governed by a highly polygenic and environment-dependent genetic architecture with widespread conditional pleiotropy and context-dependent fitness effects for genes, plasmids and clones. Our results reveal that fitness is strongly shaped by genotype–environment interactions, with substantial variation in the contribution of different genomic modalities across condition categories. Genomic associations highlight the role of well-characterized resistance and virulence determinants, clinically relevant plasmids, and previously uncharacterized but mechanistically plausible genes in shaping the bacterial fitness landscape. Furthermore, we show that both population structure and lineage-independent signals from accessory genes and SNPs provide moderate but consistent predictive power across conditions. Together, our results support a shift from considering bacterial fitness as a static genotype-encoded trait to an environment-dependent landscape shaped by interactions between genetic variation and ecological context.

### The accessory genome provides adaptive benefits but carries fitness costs, shaped by environmental trade-offs

Our study extends previous chemical genomics studies by covering a broad range of growth conditions and directly profiling a large collection of clinical isolates, rather than relying on single-gene deletion libraries under limited conditions. Traditional forward genetic platforms infer gene function by measuring the fitness effects of mutants carrying targeted gene knockouts [19]. While this provides a straightforward framework for functional inference, such approaches are inherently limited to the genetic background of the strain from which the library is derived, restricting annotation to the core genome and overlooking the accessory genome, which harbours key determinants of *K. pneumoniae* adaptation, virulence, and resistance. By integrating chemical genomics with natural strain variation, our approach enabled functional inference across the pangenome. Although lineage structure and genome-wide linkage can complicate attribution, this framework provides valuable insights into accessory gene function, many of which remain poorly characterized. Galardini et al. demonstrated the utility of high throughput profiling in *E. coli*, albeit without capturing pangenome-wide variation [59]. In contrast, our approach reveals structured relationships between genetic and phenotypic diversity across conditions, highlighting the accessory genome, including mobile elements, regulatory genes, and virulence-associated loci, as a key adaptive layer. This framework converges with transposon-based and CRISPRi fitness screens in *K. pneumoniae*, which collectively show that fitness is inherently environment-dependent, with gene essentiality emerging only under specific stresses, and shaped by strain variation, redundancy, and adaptive trade-offs within a dynamic fitness landscape [9–11, 18, 60]. Our results identify hypervirulence-associated siderophore systems, particularly aerobactin and, to a lesser extent, yersiniabactin, as key accessory determinants for fitness across diverse conditions, reflecting the metabolic cost of siderophore production and its associated fitness trade-offs outside iron-limited environments [61]. A recent study using gene-knockout mutants and mouse infection models demonstrated that the hypervirulence plasmid commonly associated with the hv ST23 clone enhances pathogenicity while imposing a measurable fitness cost, as reflected by improved growth of plasmid-cured strains and frequent plasmid loss under non-selective conditions, highlighting a trade-off between virulence and basal fitness [62]. More broadly, many bacterial virulence genes exhibit antagonistic pleiotropy, conferring benefits in specific host niches while imposing costs in alternative environments. Constitutive virulence expression imposes fitness costs, as shown by reduced growth upon PrfA activation in *L. monocytogenes* and slower growth of TTSS-1–expressing Salmonella cells [63]. In line with these findings, our results reinforce the idea that the accessory genome, particularly virulence-associated genes, confers environment-dependent benefits while imposing fitness costs, reflecting trade-offs across conditions.

### Heritability and predictability of bacterial phenotypes are environment-dependent, revealing architectural constraints across conditions

Our predictive framework extends previous efforts in genome-based bacterial phenotype prediction from a narrow set of traits to a range of traits across broad conditions. Earlier studies primarily focused on pathogen-intrinsic traits, quantitative and environmentally sensitive phenotypes [56, 64], or host-integrated clinical outcomes [65, 66], but predicted limited number of traits or conditions. By expanding the phenotypic landscape, our study provides a comprehensive view of bacterial fitness across diverse environmental niches. Our results are consistent with these prior observations. For several pathogen-intrinsic traits, e.g. antibiotic MICs for multiple drugs in *K. pneumoniae* [67], *E. coli* [68] and *S. pneumoniae* [52], bacterial genomes is known to explain a large fraction of phenotypic variance and exhibit high heritability, consistent with patterns observed for antibiotic resistance, stress combinations, and certain selective environmental stress conditions in our study. In contrast, for intermediate, quantitative, and environmentally sensitive traits (such as growth under sub-MIC stress [56], plasmid permissiveness [69], and complex virulence phenotypes [64, 70]), genomic variation explains a moderate but clearly incomplete fraction of phenotypic variance, with strong dependence on environmental context, as we also reported for a range of environmental conditions. This phenomenon, often referred to as bacterial genetic dark matter, is increasingly recognized across diverse pathogens and traits, where GWAS, heritability analyses, and machine learning models approach saturation yet leave a substantial fraction of phenotypic variance unexplained [56, 65, 71, 72]. While part of this unexplained variance can be attributed to phenotype noise and assay heterogeneity, the high reproducibility of our measurements suggests that technical noise contributes only marginally to the observed limits in predictability. Furthermore, incorporating predictive uncertainty in our setting yielded prediction intervals, which allowed us to capture variability in inherently noisy assay measurements and improve the reliability of predictions. Instead of the technical features, the low predictability and heritability in non-antibiotic environmental stress conditions likely reflect lineage-structured variation arising from epistasis, rare and accessory variation, regulatory effects, environmental context, and strong population structure. These limitations are known to be inherent to sequence-only approaches, which fail to capture regulatory, and systems-level mechanisms [70, 73]. Bridging the gap between genotype and phenotype will therefore require multi-omics predictive and association frameworks that integrate genomic data with transcriptomic and proteomic layers, explicitly model regulatory networks and gene–gene interactions [70, 73]. Such integration will enable a deeper understanding of the architecture and limits of fitness predictability across molecular layers.

### Plasmid fitness is a context-dependent landscape shaped by host and environment

Our results systematically demonstrate that plasmid fitness effects are highly variable across host genotypes and environments, challenging the notion of a single, fixed plasmid cost of carriage. Instead, we observed multidimensional variability, where plasmid effects on fitness depend on both host background and environmental context. Across conditions, this effect was broadly monotonic with respect to plasmid replicon types, but the direction varied: increasing replicon presence was associated with either increased or decreased fitness depending on whether conditions involved antibiotic stress, stress combinations, or non-antibiotic environments. In particular, plasmids exhibited mild fitness costs under non-antibiotic conditions but substantial fitness effects under selective pressures, with marked variation across lineages. Within this context, the same plasmid can be costly, neutral, or beneficial depending on context. This is consistent with prior studies in more limited settings; specifically, the pOXA-48 plasmid, carrying the carbapenemase *bla*_OXA-48_ gene, was shown to impose an overall fitness cost in many hosts but can confer neutral or even beneficial effects depending on bacterial host genetic background [74]. More broadly, previous work also demonstrated that environmental parameters could alter not only the magnitude but also the direction of plasmid fitness effects, modulating the balance between vertical inheritance and horizontal transfer [75–77].These studies were, however, typically restricted to a limited range of environments or host systems. In contrast, our study systematically characterizes plasmid-associated fitness effects across a large population and diverse conditions. Based on these findings, we frame plasmid effects within a multidimensional fitness landscape shaped by interactions between plasmid genes, host genomic background, and environment [78]. This framework highlights the adaptive flexibility of plasmids and provides a potential explanation for their long-term persistence in clinical populations through context-dependent fitness advantages rather than constant selective benefit.

### Limitations

Despite the breadth of the collection and the depth of the analysis, several limitations should be acknowledged. First, the strain collection was derived primarily from clinical settings and therefore does not fully capture the global diversity of *K. pneumoniae*. Although major clinical lineages were represented, the dataset was dominated by a limited number of clonal groups, which may bias estimates of gene effects in association analyses. In particular, the clinical origin of the samples leads to overrepresentation of specific lineages, limiting the applicability of our findings to community and environmental settings, where *K. pneumoniae* diversity was shown to differ substantially in One Health studies [79]. This limitation may restrict insights into fitness determinants relevant to non-clinical settings. Second, the predominance of multidrug-resistant and hypervirulent strains in our collection led to strong correlations between resistance and virulence determinants. While this linkage can enhance predictive performance by providing multiple correlated biomarkers, it complicates the identification of causal features and the interpretation of feature importance. Future studies incorporating broader sampling, including environmental and non-clinical strains, will be essential to provide a more comprehensive view of *K. pneumoniae* diversity, uncover novel genetic determinants of survival across diverse ecological niches, and improve the generalizability of predictive models.

### Conclusion

This work established a predictive framework for understanding bacterial fitness across clinically and non-clinically relevant environments. By integrating large-scale chemical genomics with machine learning, we showed that fitness is not a fixed property of genotype but is shaped by interactions between the genome and the environment. Quantifying the fitness effects of genes and variants provides a practical route for prioritising functional validation, which may enable lineage-informed therapeutic strategies and fitness-aware genomic surveillance in future.

## Data Availability

We developed the ColonyExplorer portal, which contains the results from the screening assays and colony characterizations. The portal allows users to select colonies based on genome accession numbers and directly visualize growth and quantified growth features. We also provide all assay results, intermediate files, GWAS and machine learning results, along with the consensus sequences of gene families from pan-genome analysis, within the portal (https://colonyexplorer.kaust.edu.sa/ and https://github.com/gzhoubioinf/ColonyExplorer). The trained models are made available as a command-line application, allowing users to input genome assemblies and predict relative fitness under selected conditions. All R and Python codes used for modelling is also available in the project's GitHub repository (https://github.com/Sara-Iftikhar/ChemGenomics). Genomic data have been submitted to GeneBank databse with accession numbers detailed in Supplemental Table S2.

https://github.com/gzhoubioinf/ColonyExplorer

## Acknowledgment

G.Z., G.W., M.T.M., R.A., W.A., O.F., M.M., S.I., and D.M. were supported by KAUST baseline funding (BAS/1/1108-01-01) and KAUST Center of Excellence for Smart Health (FCC/1/5932-01-03). M.B. was supported by a UKRI Future Leaders Fellowship (MR/V027204/1). R. A. was supported by KAIMRC funding (IRB/0026/24).

## Author Contribution

G.Z., G.W., M.T.M., M.B., R.A. and D.M. contributed equally to this work. G.Z., G.W., M.T.M., M.M., R.A., W.A., O.F., and S.I. conducted the experimental and computational research. A.H., M.R., D.M., M.B. and R.A. provided input and resources. M.B., D.M. and R.A. acquired funding. V.M., and D.M. and M.B. interpreted the results and contributed to the conceptualisation and design of the analytical framework. M.B, D.M. and R.A. supervised the study. G.Z., G.W., M.B., and D.M. wrote the manuscript with input from all authors. All authors have read and approved the manuscript.

## Ethical approvals

This study received approval from the Saudi Ministry of National Guard Health Affairs (MNGHA) (approval number NRC23R/533/08) and from the Institutional Review Board (IRB) of King Abdullah University for Science and Technology (approval number 17IBEC38). The research described here was conducted in accordance with the Helsinki declaration.

